# Premature aging in children and adults with a Fontan circulation

**DOI:** 10.1101/2023.04.25.23288924

**Authors:** Nigel E Drury, John Stickley, Thomas P Gaffey, Wei Guo, Xiaojing Yang, Yap Ching Chew, Rami Dhillon, Paul F Clift

## Abstract

The Fontan circulation predisposes to multiple morbidities typically associated with older age but occurring in their third and fourth decades, consistent with premature aging. In this pilot study, we evaluated whether the Fontan circulation is associated with premature epigenetic aging. We found that in whole blood, Δage, the difference between chronological age and estimated epigenetic DNAge™, was significantly higher in those living with a Fontan circulation than in a reference cohort (*z*=5.59, p<0.0001), with a mean Δage in adults post-Fontan of +4.9 years (95% CI 3.3-6.5, p<0.0001). Our results suggest that the Fontan circulation is associated with accelerated epigenetic aging, mirroring the premature aging clinical phenotype. In a life-limiting condition with few treatment options, targeting epigenetic aging may represent a novel opportunity for intervention.

## Introduction

The Fontan circulation represents a final common pathway for those born with a single ventricle heart condition, whereby the functioning ventricle is used to support the systemic circulation with passive pulmonary blood flow in series. However, with elevated central venous pressure and impaired cardiac output, this inherently inefficient circulation predisposes to multiple morbidities, including progressive functional decline, neurocognitive impairment, sarcopenia, osteoporosis, renal dysfunction, and liver fibrosis [1]; these clinical manifestations, typically associated with older age, are consistent with premature aging occurring in their third and fourth decades. Consequently, a 40-year-old patient with Fontan physiology has an 18% 5-year risk of death, comparable with a 75-year-old in the general population [2]. It therefore remains a life-limiting condition with limited treatment options.

Epigenetic clocks, based on the extent of DNA methylation at multiple 5’-cytosine-phosphate-guanine-3’ (CpG) sites, have been used to estimate epigenetic age, which is tightly correlated with chronological age in human controls [3]. In this pilot study, we evaluated whether the Fontan circulation is associated with premature epigenetic aging.

## Methods

### Study population

Patients on a single ventricle pathway under follow-up in Birmingham, UK were recruited, excluding any known chromosomal or genetic disorders. With ethical approval and written informed consent, clinical data and blood were obtained during a routine hospital visit.

### Reference cohort

449 healthy volunteers, without known age-related disease (median chronological age 34 years, IQR 27-49).

### Sample preparation

Whole blood samples were collected in EDTA or DNA/RNA Shield and stored at -80°C. Sample DNA was purified using the *Quick*-DNA Miniprep Plus kit (Zymo Research, Irvine, CA), and quantity/quality control checks performed. Bisulfite conversion was performed using the EZ DNA Methylation-Lightning kit (Zymo Research) according to protocol. Samples were enriched for sequencing of >2,000 age-associated gene loci using Illumina NovaSeq technology (Illumina, San Diego, CA).

### Sequence Alignments & Data Analysis

Sequence reads were identified using Illumina base calling software and aligned to the reference genome using Bismark, an aligner optimized for bisulfite sequence data and methylation calling. The methylation level of each sampled cytosine was estimated as the number of reads reporting C, divided by the total number of reads reporting C or T. Calculated DNA methylation values obtained from the sequence data were used to assess DNA methylation age (DNAge™) according to Zymo’s proprietary algorithm.

### Statistical analysis

Analysis was performed using R version 3.6. Δage was calculated as the difference between chronological age (date of sample – date of birth) and estimated DNAge™. One sample t-test was used to determine if the mean difference was equal to zero, and Mann-Whitney U-test to compare groups, with statistical significance set at p <0.05.

## Results

Blood was obtained from 42 patients: two children pre-Fontan completion (chronological age 4 years), ten children post-Fontan (median 10.3 years, IQR 9.4-13.4), and 30 adults living with a Fontan circulation (median 29.8 years, IQR 24.0-35.3). Post-Fontan, overall mean Δage was +4.4 years (95% CI 3.1-5.7, p<0.0001), in the children +2.8 years (95% CI 1.3-4.4, p=0.0025) and in the adults +4.9 years (95% CI 3.3-6.5, p<0.0001), compared with median +0.6 years (IQR-1.6-2.8) in the reference cohort. Δage was significantly higher post-Fontan than in controls (*z*=5.59, p<0.0001), and in males versus females with a Fontan circulation (*z*=2.40, p=0.016). Correlations between DNAge and chronological age are shown in the figure. Only three patients post-Fontan had a negative Δage, all of whom had a chronological age >35 years.

**Figure.**
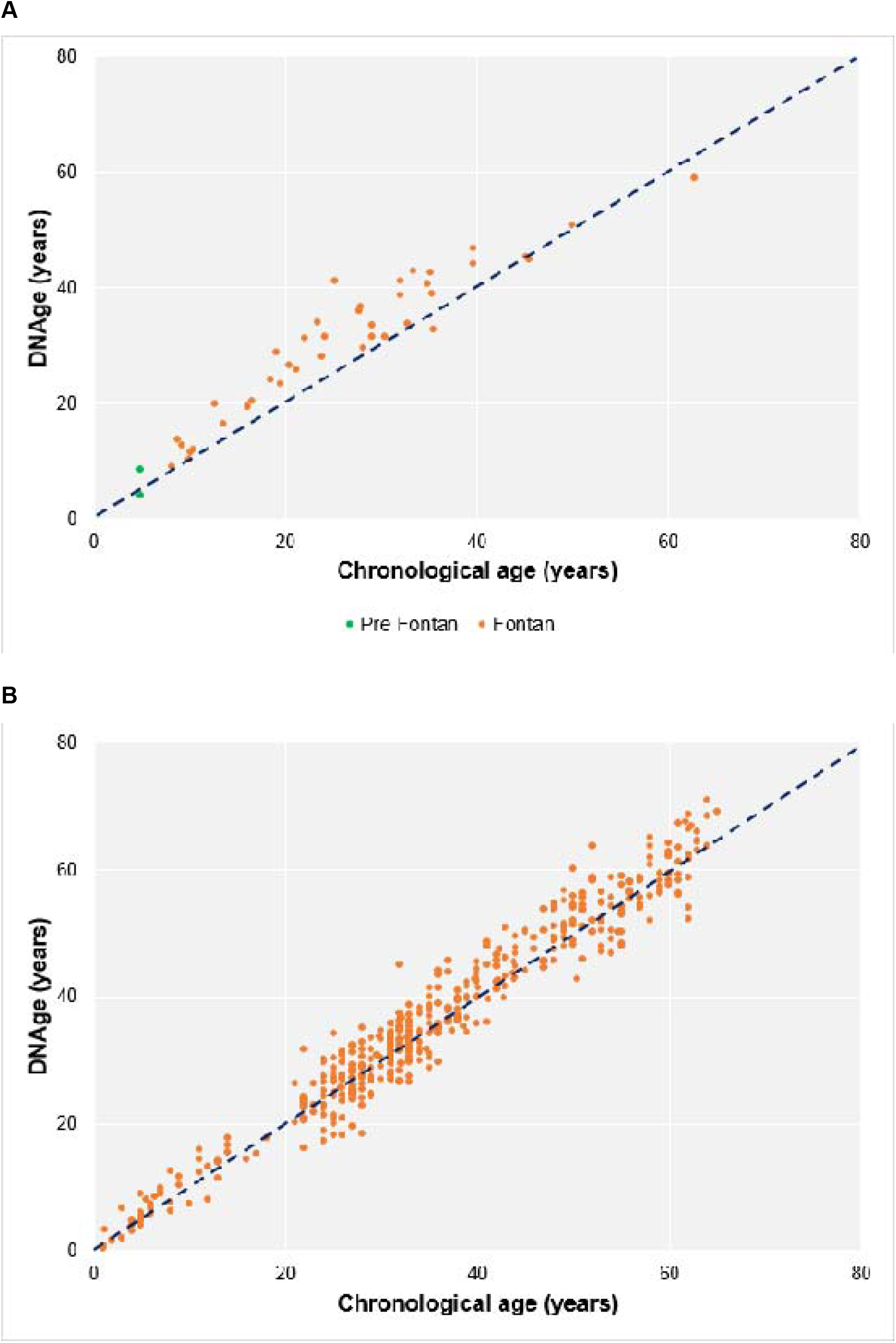
Correlation of chronological age at the time of blood sampling with estimated DNAge in A) the patient cohort, and B) the reference cohort.

## Discussion

This study is the first to demonstrate that the Fontan circulation is associated with accelerated epigenetic aging, mirroring the premature aging clinical phenotype. Our findings suggest that this process occurs from childhood and continues during adult life, potentially due to the chronic systemic stress of Fontan physiology. The mean Δage of +4.9 years in our adult cohort is consistent with the extent of premature epigenetic aging seen in other multi-system conditions associated with an accelerated aging phenotype, such as adults living with chronic human immunodeficiency virus (HIV) infection (mean +5.2 years) [4].

As a pilot study, we had insufficient data to determine the relationship between chronological age and Δage; the equivalence seen in the older patients may represent sampling bias or survivorship bias in those undergoing Fontan completion over 30 years ago when late survival was rare. We were also unable to explore the relationship between DNAge and morphological or clinical factors, such as systemic ventricle, type of Fontan, atrial fenestration, or clinical status; evaluating these variables in a larger clinical-epigenetic dataset will provide insight into their relative impact on the premature aging profile.

As a surgically constructed palliation during early childhood, in which there are currently limited options to improve durability during adulthood, targeting epigenetic aging may represent a novel treatment opportunity [5], to slow down or halt disease progression. The effects of pharmacological therapies, physical exercise, and nutritional strategies on premature aging in patients living with a Fontan circulation are unknown.

## Data Availability

All data produced in the present study are available upon reasonable request to the authors.

## Acknowledgements

We thank Prof Janet M Lord, University of Birmingham, and Dr Leda Mirbahai, University of Warwick, for their advice on accelerated aging and epigenetic profiling. We thank the patients who generously gave blood for the study, and gratefully acknowledge the contribution to this study made by the University of Birmingham’s Human Biomaterials Resource Centre which has been supported through Birmingham Science City – Experimental Medicine Network of Excellence project. Ethical approval was obtained from the North West – Haydock NHS Research Ethics Committee (20/NW/0001) on 20^th^ February 2020.

## Sources of Funding

This work was funded by a generous donation from the Charlie Ramsey Research Fund to the Birmingham Children’s Hospital Charity (37-6-175). Nigel Drury was funded by an Intermediate Clinical Research Fellowship from the British Heart Foundation (FS/15/49/31612).

## Disclosures

WG, XY and YCC are employees of Zymo Research Corporation. All other authors have no conflicts of interest to disclose.

## Notes

### Funding Statement

This study was funded by a generous donation from the Charlie Ramsey Research Fund to the Birmingham Childrens Hospital Charity (37-6-175). Nigel Drury was funded by an Intermediate Clinical Research Fellowship from the British Heart Foundation (FS/15/49/31612).

### Author Declarations

The North West-Haydock NHS Research Ethics Committee gave ethical approval for this work via application ref: 20/NW/0001 on 20 February 2020.

## References

1. Rychik J, Goldberg DJ. Late consequences of the Fontan operation. Circulation 2014;130:1525–8.

2. Diller GP, Kempny A, Alonso-Gonzalez R, Swan L, Uebing A, Li W, Babu-Narayan S, Wort SJ, Dimopoulos K, Gatzoulis MA. Survival prospects and circumstances of death in contemporary adult congenital heart disease patients under follow-up at a large tertiary centre. Circulation 2015;132:2118–25.

3. Horvath S. DNA methylation age of human tissues and cell types. Genome Biol 2013;14:R115.

4. Horvath S, Levine AJ. HIV-1 infection accelerates age according to the epigenetic clock. J Infect Dis 2015;212(10):1563–73.

5. Fahy GM, Brooke RT, Watson JP, Good Z, Vasanawala SS, Maecker H, Leipold MD, Lin DTS, Kobor MS, Horvath S. Reversal of epigenetic aging and immunosenescent trends in humans. Aging Cell 2019;18:e13028.

